# Associations between e-cigarette marketing exposure and vaping nicotine and cannabis among U.S. adults, 2021

**DOI:** 10.1101/2024.02.03.24302079

**Authors:** Julia Chen-Sankey, Kathryn La Cparia, Allison Glasser, Alisa A. Padon, Meghan B. Moran, Kimberly G. Wagoner, Kristina M. Jackson, Carla J. Berg

## Abstract

**Importance:** Vaping has become an increasingly common method for consuming nicotine and cannabis, a trend potentially influenced by e-cigarette marketing. However, little is known about the influence of e-cigarette marketing on cannabis vaping behaviors.

**Objective:** To examine the associations between e-cigarette marketing exposure and nicotine and cannabis vaping behaviors among adults.

**Design, Setting, and Participants:** This cross-sectional study included a U.S. nationally representative sample of adults (≥18 years) from the Wave 6 survey of the Population Assessment of Tobacco and Health (PATH) Study, conducted from March to November 2021.

**Exposure:** Past 30-day e-cigarette marketing exposure (overall and by ten marketing channels).

**Main Outcomes and Measures:** Past 30-day vaping behavior (sole- and dual-vaping of nicotine and cannabis) overall and stratified by age.

**Results:** The study included 30,516 respondents (48.0% male and 63.9% non-Hispanic White). Overall, 52.0% of respondents reported past 30-day e-cigarette marketing exposure, and 89.8%, 5.6%, 3.2%, and 1.4% reported no vaping, sole-nicotine vaping, sole-cannabis vaping, and dual-vaping, respectively. Multinominal logistic regression results show exposure to e-cigarette marketing was associated with increased odds of reporting sole-cannabis vaping versus no vaping (adjusted risk ratio [aRR], 1.31; 95% confidence interval [CI], 1.09-1.57) and dual-vaping versus no vaping (aRR, 1.26; 95% CI, 1.01-1.57). Stratification analysis found these associations among those aged 18-24 and 25-34 years but not older adults (≥35 years). Those exposed to e-cigarette marketing also had increased odds of reporting sole-cannabis vaping versus sole-nicotine vaping (aRR, 1.28; 95% CI, 1.04-1.58). Stratification analysis found this association only among those aged 18-24 years. E-cigarette marketing exposure via several channels (retail stores, billboards, events, newspapers/magazines) was associated with increased odds of reporting sole-cannabis vaping.

**Conclusions and Relevance:** E-cigarette marketing exposure was only associated with sole-cannabis vaping and dual-vaping, not sole-nicotine vaping among U.S. adults. Such associations were mainly driven by young adults aged 18-24 and 25-35 years and were found for multiple marketing channels. Greater restrictions on tobacco marketing may have reduced the influence of e-cigarette marketing on nicotine vaping, while gaps in such marketing restrictions for cannabis may contribute to continued influence of e-cigarette marketing on cannabis vaping.

**KEY POINTS:** *Question:* What is the association between e-cigarette marketing exposure and nicotine and cannabis vaping behaviors among U.S. adults?

*Findings:* In this cross-sectional study of 30,516 adults, those exposed to e-cigarette marketing were about 1.3 times more likely to report sole-cannabis vaping and dual-nicotine and cannabis vaping compared to those not exposed to e-cigarette marketing. Such associations were not found for sole-nicotine vaping.

*Meaning:* Greater restrictions on tobacco marketing may have reduced the influence of e-cigarette marketing on nicotine vaping, while gaps in marketing restrictions for cannabis may contribute to the continued influence of e-cigarette marketing on cannabis vaping.

## INTRODUCTION

In recent years, e-cigarette or vape products with nicotine, have become one of the most commonly used tobacco products in the U.S. The prevalence of past 30-day e-cigarette use with nicotine (or vaping nicotine) was 17.0% among middle and high school-aged youth in 2023^1^ and 17.2% and 5.2% among young adults (ages 19-30) and midlife adults (ages 35-50), respectively, in 2022.^2^ During the same time, many states legalized non-medical (or recreational) cannabis retail, coinciding with increased vape product use with cannabis (often called vaping cannabis or vape pens). In 2022, 13.9% and 6.3% of U.S. young adults and midlife adults reported past 30-day cannabis vaping, respectively.^2^ The share of e-cigarettes or vape products in the cannabis market has also expanded—ranking second in 2023, after flower products.^3^ Overall, vaping has become an increasingly common consumption method for both nicotine and cannabis.

Mounting evidence has demonstrated that e-cigarette marketing might be one of the main contributing factors to the uptake and continued behavior of vaping nicotine across ages.^4–7^ In contrast, there is limited understanding of whether e-cigarette marketing exposure is associated with vaping cannabis, including vaping only cannabis and vaping nicotine and cannabis concomitantly.^8^ Previous studies that examined the influence of cannabis product marketing only focused on the marketing of all cannabis products without specifically examining cannabis vape product marketing or use behaviors.^9–14^ Additionally, those studies often examined cannabis marketing presence and exposure at dispensaries or retailers or used convenience samples of participants who did not represent the overall nationwide population. Lastly, these studies rarely investigated such associations by marketing exposure channels (e.g., retailers, billboards, magazines/newspapers) or the population’s age groups. E-cigarette marketing exposure through certain channels may be associated with vaping cannabis more than through other channels; it is also likely that such associations only exist or are stronger among younger rather than older adult populations.^15^

This study used secondary data from the Population Assessment of Tobacco and Health (PATH) Study adult survey^16,17^ to address these research gaps and inform our understanding of the associations of e-cigarette marketing exposure with vaping nicotine and cannabis. Specifically, the study examined cross-sectional associations between e-cigarette marketing exposure (by marketing channels) and vaping behaviors categorized by vaping nicotine, cannabis, or both among U.S. adults, stratified by age.

## METHODS

### Study Sample

We used data from Wave 6 adult survey public-use files of the PATH Study, which includes nationally representative, longitudinal cohorts of civilian, noninstitutionalized individuals in the U.S.^16^ In-person data collection and telephone data collection procedures were used for Wave 6 and occurred between March and November 2021. Adults who completed the interview received $50. More details about the PATH Study can be found elsewhere.^16,17^ All respondents from the adult survey were included in the analysis, resulting in a sample size of *n*=30,516. This study was exempted from Rutgers University institutional review board approval because it involves the use of publicly available data whereby subjects cannot be identified. The study followed the Strengthening the Reporting of Observational Studies in Epidemiology (STROBE) reporting guideline for cohort studies.^18^

#### Past 30-Day E-cigarette Marketing Exposure

At Wave 6, respondents were asked: “In the past 30 days, have you noticed e-cigarettes or other electronic nicotine products being advertised in any of the following places?” (“Yes” and “No” options were displayed for each of the following channels): “Gas stations, convenience stores, or other retail stores,” “On billboards,” “In newspapers or magazines,” “On radio,” “On television,” “At events like fairs, festivals, or sporting events,” “At nightclubs, bars, or music concerts,” “In email messages,” “On social media,” “On other websites online,” and “Somewhere else.” We created an overall binary variable that indicated any e-cigarette marketing exposure via any channel versus no exposure as well as ten variables based on e-cigarette marketing exposure versus no exposure via each channel.^4,5,7^

#### Past 30-Day Vaping Behavior

Respondents were asked: “In the past 30 days, have you ever used an electronic nicotine product, even once or two times? Please do not include marijuana or cannabis when answering the following questions about electronic nicotine products.” A variable was created to reflect any past 30-day nicotine vaping (Weighted %=6.9%) versus none. Respondents were also asked about their use of marijuana (defined by the PATH Study as cannabis, pot, weed, THC, hash, kush, or CBD that includes smoked, vaped, or ingested types of marijuana or cannabis).^16^ Those who reported past 30-day marijuana use were further asked, “In the past 30 days, which of the following ways did you use marijuana? Choose all that apply.” Those who chose “Vape marijuana liquids or oils in an e-cigarette, vape pen, or electronic nicotine product” were considered reporting past 30-day cannabis vaping (Weighted %=4.6%) versus none. We then used past 30-day nicotine and cannabis vaping measures to create a new variable that captures four types of vaping behavior: (1) no vaping (having not vaped nicotine or cannabis), (2) sole-nicotine vaping (vaped only nicotine), (3) sole-cannabis vaping (vaped only cannabis), and (4) dual-vaping (vaped both nicotine and cannabis).^19^

### Covariates

Covariates included sociodemographic characteristics: age, biological sex, race and ethnicity, sexual orientation, and annual household income (see Table 1 for variable categories). Race and ethnicity were included as social constructs instead of biological or generic categories. We also included self-rated physical and mental health statuses as well as past 30-day use of other cannabis products (smoked dried herb or flower or other ways) and other tobacco products (smoked tobacco, or used hookah, snus, smokeless tobacco etc.). Age (18-24, 25-34, 35-54, and ≥55 years)^20–22^ was also used as a stratification variable for regression models.

**Table 1.** Participant characteristics overall and by past 30-day e-cigarette marketing exposure^a-d^.

|  |  | Past 30-Day E-cigarette Marketing Exposure |  |  |  |  |  | P-value |
| --- | --- | --- | --- | --- | --- | --- | --- | --- |
|  |  | All respondents |  | Exposed |  | Not Exposed |  |  |
|  |  | % | 95% CI | % | 95% CI | % | 95% CI |  |
| Overall |  |  |  | 52.0 | 51.0, 53.0 | 48.0 | 47.3, 49.1 |  |
| Age |  |  |  |  |  |  |  | <0.001 |
|  | 18-24 | 11.6 | 11.5, 11.8 | 13.6 | 13.2, 13.9 | 9.7 | 9.3, 10.0 |  |
|  | 25-34 | 16.9 | 16.4, 17.3 | 18.8 | 18.2, 19.6 | 14.7 | 14.0, 15.4 |  |
|  | 35-54 | 31.7 | 31.1, 32.4 | 33.0 | 31.9, 34.1 | 30.3 | 29.2, 31.5 |  |
|  | ≥55 | 39.8 | 39.2, 40.3 | 34.6 | 33.5, 35.8 | 45.3 | 44.2, 46.5 |  |
| Biological Sex |  |  |  |  |  |  |  | 0.003 |
|  | Female | 52.0 | 51.8, 52.3 | 50.6 | 49.7, 51.6 | 53.5 | 52.5, 54.6 |  |
|  | Male | 48.0 | 47.8, 48.2 | 49.4 | 48.5, 50.3 | 46.5 | 45.4, 47.5 |  |
| Race and Ethnicity |  |  |  |  |  |  |  | 0.005 |
|  | Non-Hispanic White | 63.9 | 63.7, 64.1 | 64.5 | 63.6, 65.3 | 63.3 | 62.3, 64.4 |  |
|  | Non-Hispanic Black | 11.4 | 11.2, 11.5 | 11.8 | 11.4, 12.2 | 10.9 | 10.4, 11.4 |  |
|  | Hispanic | 16.5 | 16.3, 16.7 | 16.3 | 15.6, 17.1 | 16.7 | 15.9, 17.5 |  |
|  | Non-Hispanic Other | 8.2 | 8.1, 8.4 | 7.4 | 6.9, 8.0 | 9.1 | 8.6, 9.7 |  |
| Sexual Orientation |  |  |  |  |  |  |  | <0.001 |
|  | Heterosexual | 92.4 | 92.0, 92.8 | 91.4 | 90.8, 91.9 | 93.6 | 92.9, 94.2 |  |
|  | Non-heterosexual | 7.6 | 7.2, 8.0 | 8.6 | 8.1, 9.2 | 6.4 | 5.8, 7.1 |  |
| Annual Household Income |  |  |  |  |  |  |  | <0.001 |
|  | <10,000 | 7.6 | 7.2, 8.2 | 6.7 | 6.2, 7.3 | 8.7 | 7.9, 9.5 |  |
|  | 10,000-24,999 | 13.9 | 13.3, 14.5 | 13.6 | 12.8, 14.5 | 14.2 | 13.3, 15.1 |  |
|  | 25,000-49,999 | 20.7 | 19.9, 21.5 | 21.3 | 20.2, 22.5 | 19.9 | 18.7, 21.2 |  |
|  | 50,000-99,999 | 27.1 | 26.2, 28.1 | 28.9 | 27.6, 30.1 | 25.2 | 24.0, 26.6 |  |
|  | ≥100,000 | 25.4 | 24.5, 26.3 | 25.7 | 24.6, 26.9 | 25.0 | 23.7, 26.4 |  |
|  | Undetermined | 5.3 | 4.9, 5.8 | 3.8 | 3.4, 4.2 | 7.0 | 6.2, 7.8 |  |
| Physical Health |  |  |  |  |  |  |  | <0.001 |
|  | Excellent | 17.9 | 17.3, 18.6 | 16.4 | 15.5, 17.4 | 19.6 | 18.5, 20.7 |  |
|  | Very good | 37.8 | 36.8, 38.7 | 38.9 | 37.6, 40.3 | 36.5 | 35.1, 37.9 |  |
|  | Good | 30.5 | 29.6, 31.4 | 31.1 | 29.8, 32.3 | 29.9 | 28.7, 31.1 |  |
|  | Fair | 11.6 | 10.9, 12.3 | 11.6 | 10.8, 12.4 | 11.6 | 10.6, 12.8 |  |
|  | Poor | 2.2 | 2.0, 2.5 | 2.1 | 1.8, 2.4 | 2.4 | 2.1, 2.9 |  |
| Mental Health |  |  |  |  |  |  |  | <0.001 |
|  | Excellent | 20.2 | 19.5, 20.9 | 18.3 | 17.3, 19.2 | 22.2 | 21.2, 23.4 |  |
|  | Very good | 35.3 | 34.4, 36.3 | 35.5 | 34.2, 36.8 | 35.2 | 33.9, 36.6 |  |
|  | Good | 29.0 | 28.2, 29.8 | 29.3 | 28.1, 30.4 | 28.7 | 27.5, 29.9 |  |
|  | Fair | 12.7 | 12.1, 13.3 | 14.1 | 13.3, 14.9 | 11.2 | 10.4, 12.1 |  |
|  | Poor | 2.8 | 2.5, 3.0 | 2.9 | 2.6, 3.3 | 2.6 | 2.3, 2.9 |  |
| Past 30-Day Other Cannabis Product Use |  |  |  |  |  |  |  | <0.001 |
|  | No | 85.4 | 84.8, 86.0 | 83.5 | 82.7, 84.2 | 87.4 | 86.6, 88.2 |  |
|  | Yes | 14.6 | 14.0, 15.3 | 16.5 | 15.8, 17.3 | 12.6 | 11.8, 13.4 |  |
| Past 30-Day Other Tobacco Product Use |  |  |  |  |  |  |  | 0.001 |
|  | No | 79.1 | 78.5, 79.7 | 78.1 | 77.3, 78.8 | 80.2 | 79.2, 81.0 |  |
|  | Yes | 20.9 | 20.3, 21.5 | 21.9 | 21.2, 22.7 | 19.8 | 19.0, 20.8 |  |
| Past 30-Day Vaping Behavior |  |  |  |  |  |  |  | <0.001 |
|  | No vaping | 89.8 | 89.5, 90.2 | 88.2 | 87.7, 88.8 | 91.6 | 91.0, 92.1 |  |
|  | Sole-nicotine vaping | 5.6 | 5.3, 5.8 | 6.1 | 5.7, 6.5 | 5.0 | 4.7, 5.4 |  |
|  | Sole-cannabis vaping | 3.2 | 3.0, 3.5 | 4.0 | 3.6, 4.3 | 2.4 | 2.1, 2.9 |  |
|  | Dual-vaping | 1.4 | 1.3, 1.5 | 1.7 | 1.5, 1.9 | 1.0 | 0.9, 1.2 |  |

Abbreviations: CI, confidence interval.
a. All percentages are weighted.
b. P-values are from Chi-square test results.
c. Race and ethnicity were self-identified by respondents and were based on the classifications developed by the U.S. Census Bureau (e.g., Hispanic, non-Hispanic Black, non-Hispanic White, and non-Hispanic other [including American Indian or Alaska Native, Asian, Native Hawaiian or other Pacific Islander, and more than 1 race]).
d. Non-heterosexual category includes lesbian, gay, bisexual, and something else.

### Statistical Analysis

Data analyses were conducted between September and November 2023. After descriptive analyses, we estimated weighted past 30-day prevalence of e-cigarette marketing exposure (by marketing channel) and vaping behaviors overall and stratified by age. We used Chi-square tests to examine the differences between covariate variables and e-cigarette marketing exposure. In multinomial logistic regression analyses, we used separate regressions to estimate associations between past 30-day e-cigarette marketing exposure and past 30-day vaping behaviors overall and stratified by age. We also examined any past 30-day e-cigarette marketing exposure and exposure by ten channels using separate regressions overall and stratified by age. For all above-mentioned regression analyses, we used both no vaping and sole-nicotine vaping as reference groups, allowing us to detect how e-cigarette marketing exposure is related to the probability of being in the outcome group (sole-cannabis vaping and dual-vaping) versus the reference group (no vaping and sole-nicotine vaping).^23^ All regression models controlled for covariates.

We applied the Wave 6 single wave weights for the Wave 4 Cohort when calculating proportions with 95% confidence intervals (CI), adopting the balanced repeated replications method with a Fay adjustment of 0.3.^16^ It was recommended by the PATH Study that weights developed for the Wave 4 Cohort should be used for cross-sectional estimation for follow-up waves until a new cohort is established.^16^ To minimize missing data, we used imputed demographic variables and derived tobacco use variables included in the PATH public use data files and included an “undetermined” category for variables with missing values larger than 5% of the sample.^4,5,7^ For regression models, we used listwise deletion since missing data were minimal across variables used for analysis (<0.5%).^24^ Adjusted risk ratios (aRR) and 95% CIs were reported in regression analyses. Statistical analyses were performed using Stata 18.0 (Stata Corp, College Station, TX), and 2-sided *P* values <0.05 were considered statistically significant.

## RESULTS

The sample of U.S. adults was balanced in terms of sex (female: 52.0%; male: 48.0% [**Table 1**]). In terms of age, 11.7%, 16.9%, 31.7%, and 39.8% were between 18-24, 25-34, 35-54, and ≥55, respectively. In terms of race and ethnicity, 63.9%, 11.4%, 16.5%, and 8.2% self-reported non-Hispanic White, non-Hispanic Black, Hispanic, and non-Hispanic Other, respectively. Additionally, 14.6% and 20.9% of the respondents reported past 30-day use of other cannabis and tobacco products, respectively.

Overall, 52.0% of the respondents (∼134 million U.S. adults) reported past 30-day e-cigarette marketing exposure (**Table 1**). Select bivariate findings include positive associations between exposure and being 18-24 and 25-34 and reporting other cannabis or tobacco product use and vaping behaviors. **Supplemental Table 1** presents the weighted prevalence of past 30-day e-cigarette marketing exposure (by marketing channel) stratified by age and vaping behavior.

Overall, 89.8%, 5.6%, 3.2%, and 1.4% reported no vaping, sole-nicotine vaping, sole-cannabis vaping, and dual-vaping, respectively (**Table 2**). Adults aged 18-24 years and 25-34 years were more likely to report sole-nicotine vaping, sole-cannabis vaping, and dual-vaping than those aged 35-55 years and 55 years and above.

**Table 2.**
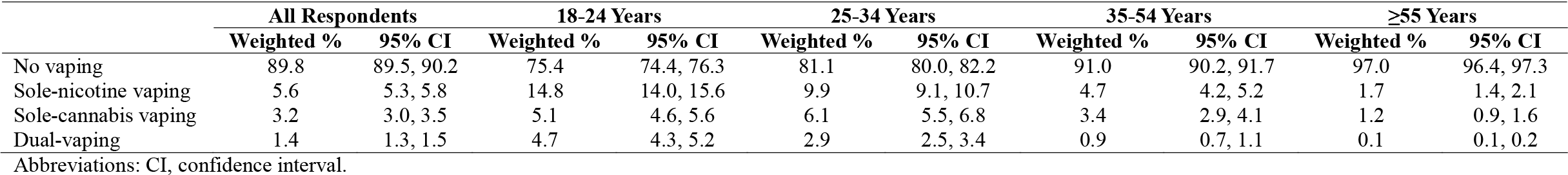
Weighted prevalence of vaping behavior in the past 30 days overall and by age group.

Past 30-day e-cigarette marketing exposure was 51.1%, 57.1%, 63.8%, and 64.9% among those who reported no vaping, sole-nicotine vaping, sole-cannabis vaping, and dual-vaping, respectively (**Table 3**). Respondents were most likely to be exposed to e-cigarette marketing through retail stores (42.2%), television (10.9%), and social media (9.3%). Compared to those who reported no vaping, those who reported sole-cannabis vaping were more likely to be exposed to e-cigarette marketing overall and through six out of the 10 marketing channels; they were also more likely to be exposed through three out of the 10 marketing channels compared to those who reported sole-nicotine vaping. Those who reported dual-vaping were more likely to be exposed to e-cigarette marketing overall and through five or three marketing channels compared to those who reported no vaping and sole-nicotine vaping, respectively.

**Table 3.**
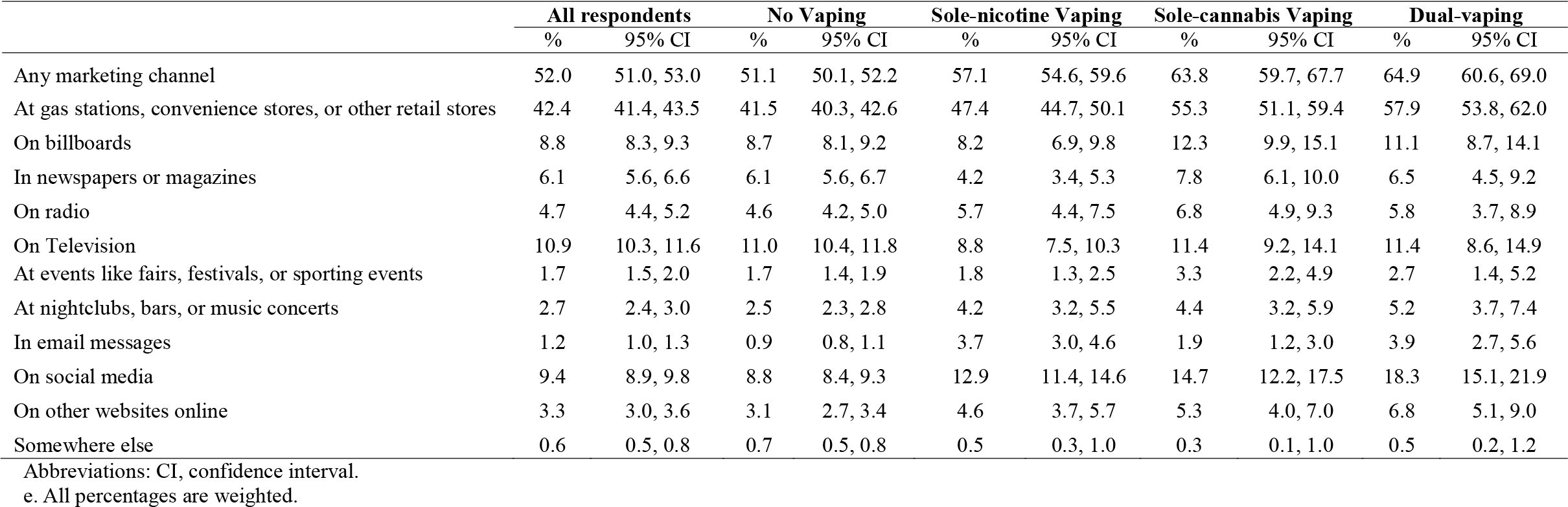
Weighted prevalence of past 30-day e-cigarette marketing exposure (by marketing channel) overall and by past 30-day vaping behavior^a^.

|  | **All respondents** |  | **No Vaping** |  | **Sole-nicotine Vaping** |  | **Sole-cannabis Vaping** |  | **Dual-vaping** |  |
| --- | --- | --- | --- | --- | --- | --- | --- | --- | --- | --- |
|  | % | 95% CI | % | 95% CI | % | 95% CI | % | 95% CI | % | 95% CI |
| Any marketing channel | 52.0 | 51.0, 53.0 | 51.1 | 50.1, 52.2 | 57.1 | 54.6, 59.6 | 63.8 | 59.7, 67.7 | 64.9 | 60.6, 69.0 |
| At gas stations, convenience stores, or other retail stores | 42.4 | 41.4, 43.5 | 41.5 | 40.3, 42.6 | 47.4 | 44.7, 50.1 | 55.3 | 51.1, 59.4 | 57.9 | 53.8, 62.0 |
| On billboards | 8.8 | 8.3, 9.3 | 8.7 | 8.1, 9.2 | 8.2 | 6.9, 9.8 | 12.3 | 9.9, 15.1 | 11.1 | 8.7, 14.1 |
| In newspapers or magazines | 6.1 | 5.6, 6.6 | 6.1 | 5.6, 6.7 | 4.2 | 3.4, 5.3 | 7.8 | 6.1, 10.0 | 6.5 | 4.5, 9.2 |
| On radio | 4.7 | 4.4, 5.2 | 4.6 | 4.2, 5.0 | 5.7 | 4.4, 7.5 | 6.8 | 4.9, 9.3 | 5.8 | 3.7, 8.9 |
| On Television | 10.9 | 10.3, 11.6 | 11.0 | 10.4, 11.8 | 8.8 | 7.5, 10.3 | 11.4 | 9.2, 14.1 | 11.4 | 8.6, 14.9 |
| At events like fairs, festivals, or sporting events | 1.7 | 1.5, 2.0 | 1.7 | 1.4, 1.9 | 1.8 | 1.3, 2.5 | 3.3 | 2.2, 4.9 | 2.7 | 1.4, 5.2 |
| At nightclubs, bars, or music concerts | 2.7 | 2.4, 3.0 | 2.5 | 2.3, 2.8 | 4.2 | 3.2, 5.5 | 4.4 | 3.2, 5.9 | 5.2 | 3.7, 7.4 |
| In email messages | 1.2 | 1.0, 1.3 | 0.9 | 0.8, 1.1 | 3.7 | 3.0, 4.6 | 1.9 | 1.2, 3.0 | 3.9 | 2.7, 5.6 |
| On social media | 9.4 | 8.9, 9.8 | 8.8 | 8.4, 9.3 | 12.9 | 11.4, 14.6 | 14.7 | 12.2, 17.5 | 18.3 | 15.1, 21.9 |
| On other websites online | 3.3 | 3.0, 3.6 | 3.1 | 2.7, 3.4 | 4.6 | 3.7, 5.7 | 5.3 | 4.0, 7.0 | 6.8 | 5.1, 9.0 |
| Somewhere else | 0.6 | 0.5, 0.8 | 0.7 | 0.5, 0.8 | 0.5 | 0.3, 1.0 | 0.3 | 0.1, 1.0 | 0.5 | 0.2, 1.2 |

Abbreviations: CI, confidence interval.
e. All percentages are weighted.

**Table 4** portrays the results from the multinomial logistic regressions. Using no vaping as the reference group, compared with adults who were not exposed to e-cigarette marketing, those who were exposed to any marketing had increased odds of reporting sole-cannabis vaping (aRR, 1.31 [95% CI, 1.09-1.57]) and dual-vaping (aRR, 1.26 [95% CI, 1.01-1.57]). Age stratification analysis showed that adults aged 18-24 years who were exposed (versus unexposed) had increased odds of sole-cannabis vaping (aRR, 1.48 [95% CI, 1.17-1.88]), and adults aged 25-34 years who were exposed (versus unexposed) had increased odds of reporting sole-cannabis vaping (aRR, 1.37 [95% CI, 1.02-1.84]) or dual-vaping (aRR, 1.46 [95% CI, 1.04-2.07]). No associations were found for those aged 35-54 years. E-cigarette marketing exposure was not associated with sole-nicotine vaping overall or among any age groups.

**Table 4.** Associations between past 30-day e-cigarette marketing exposure and past 30-day vaping behavior overall and by age^a-d^.

|  | Exposed | Not Exposed | Model 1: No vaping as the reference group | Model 2: Sole-nicotine vaping as the reference group | Exposed | Not Exposed | Model 1: No vaping as the reference group | Model 2: Sole-nicotine vaping as the reference group |
| --- | --- | --- | --- | --- | --- | --- | --- | --- |
|  | Weighted % (95% CI) | Weighted % (95% CI) | Adjusted RR (95% CI) | Adjusted RR (95% CI) | Weighted % (95% CI) | Weighted % (95% CI) | Adjusted RR (95% CI) | Adjusted RR (95% CI) |
| **All Adults** |  |  |  |  | **Adults Aged 18-24 Years** |  |  |  |
| No vaping | 88.3 (87.7, 88.8) | 91.6 (91.1, 92.1) | Reference | 0.98 (0.87, 1.09) | 75.0 (73.7, 76.2) | 76.1 (74.5, 77.5) | Reference | 1.14 (0.98, 1.34) |
| Sole-nicotine vaping | 6.1 (5.7, 6.5) | 4.9 (4.6, 5.4) | 1.02 (0.91, 1.14) | Reference | 14.1 (13.1, 15.1) | 15.6 (14.3, 17.1) | 0.87 (0.75, 1.02) | Reference |
| Sole-cannabis vaping | 4.0 (3.6, 4.3) | 2.4 (2.1, 2.9) | **1.31 (1.09, 1.57)** | **1.28 (1.04, 1.58)** | 6.0 (5.5, 6.6) | 3.7 (3.1, 4.6) | **1.48 (1.17, 1.88)** | **1.70 (1.29, 2.24)** |
| Dual-vaping | 1.6 (1.5, 1.9) | 1.1 (0.9, 1.2) | **1.26 (1.01, 1.57)** | 1.23 (0.98, 1.55) | 4.9 (4.4, 5.6) | 4.6 (3.9, 5.4) | 0.95 (0.74, 1.23) | 1.09 (0.84, 1.41) |
| **Adults Aged 25-34 Years** |  |  |  |  | **Adults Aged 35-54 Years** |  |  |  |
| No vaping | 79.7 (78.1, 81.2) | 83.3 (81.7, 84.8) | Reference | 0.86 (0.72, 1.03) | 90.0 (88.9, 91.0) | 92.1 (91.0, 93.1) | Reference | 0.99 (0.79, 1.24) |
| Sole-nicotine vaping | 10.0 (9.0, 11.0) | 9.5 (8.3, 10.7) | 1.16 (0.97, 1.39) | Reference | 5.0 (4.3, 5.7) | 4.4 (3.7, 5.2) | 1.01 (0.81, 1.26) | Reference |
| Sole-cannabis vaping | 7.0 (6.1, 8.1) | 4.9 (4.0, 6.0) | **1.37 (1.02, 1.84)** | 1.18 (0.89, 1.57) | 4.0 (3.3, 4.7) | 2.9 (2.2, 3.8) | 1.30 (0.89, 1.89) | 1.29 (0.82, 2.01) |
| Dual-vaping | 3.3 (2.8, 4.0) | 2.3 (1.8, 3.0) | **1.46 (1.04, 2.07)** | 1.26 (0.86, 1.84) | 1.0 (0.8, 1.4) | 0.6 (3.5, 1.1) | 1.52 (0.73, 3.18) | 1.51 (0.73, 3.11) |
| **Adults Aged ≥55 Years** |  |  |  |  |  |  |  |  |
| No vaping | 96.5 (95.9, 97.0) | 97.3 (96.6, 97.8) |  |  |  |  |  |  |
| Sole-nicotine vaping | 1.8 (1.5, 2.4) | 1.6 (1.2, 2.1) | NA | NA |  |  |  |  |
| Sole-cannabis vaping | 1.5 (1.1, 1.9) | 1.1 (0.6, 1.7) |  |  |  |  |  |  |
| Dual-vaping | 0.2 (0.1, 0.3) | 0.0 (0.0, 0.0) |  |  |  |  |  |  |

Abbreviations: RR, risk ratio. CI, confidence interval. NA, not available.
- Model 1 used no vaping as the reference group for the vaping behavior outcome in the multinomial logistic regressions; Model 2 used sole-nicotine vaping as the reference group for the vaping behavior outcome in the multinomial logistic regressions. - The regression models controlled for age, biological sex, race and ethnicity, annual household income, sexual orientation, physical health, mental health, past 30-day use of other cannabis products, and past 30-day use of other tobacco products. Other cannabis products: smoke dried herb or flower in a joint, pipe, hookah, or bong; Smoke dried herb or flower in a blunt cigar, cigarillo, or filtered cigar; and use marijuana some other way. Other tobacco products: cigarettes, cigarillos, filtered cigars, large cigars, pipe tobacco, hookah, snus, smokeless tobacco, and heated tobacco. - Statistically significant associations are bolded. - Results from the regression models for adults aged ≥55 years were omitted due to model convergence issues from small sample size.

Using sole-nicotine vaping as the reference group (**Table 4**), adults who were exposed (versus unexposed) to e-cigarette marketing had increased odds of sole-cannabis vaping (aRR, 1.28 [95% CI, 1.04-1.58]). Age stratification analysis showed that adults aged 18-24 years who were exposed (versus unexposed) had increased odds of sole-cannabis vaping (aRR, 1.70 [95% CI, 1.29-2.24]). No associations were found for those aged 25-34 years or 35-54 years. Regression models for those ages 55 and above did not converge due to small sample size.

Additionally, in terms of specific marketing channels, using no vaping as the reference group (**Table 5**), adults exposed to e-cigarette marketing through six channels (retail stores; billboards; newspapers or magazines; events; email messages, social media) had increased odds of sole-cannabis vaping (aRRs range between 1.30 and 1.91). Those exposed to e-cigarette marketing through email messages had increased odds of sole-nicotine vaping (adjusted RRs, 2.95 [95% CI, 2.12-4.12]). Additionally, those exposed to e-cigarette marketing through four channels (retail stores; email messages; social media; websites) had increased odds of dual-vaping (aRRs range between 1.38 and 2.97).

**Table 5.**
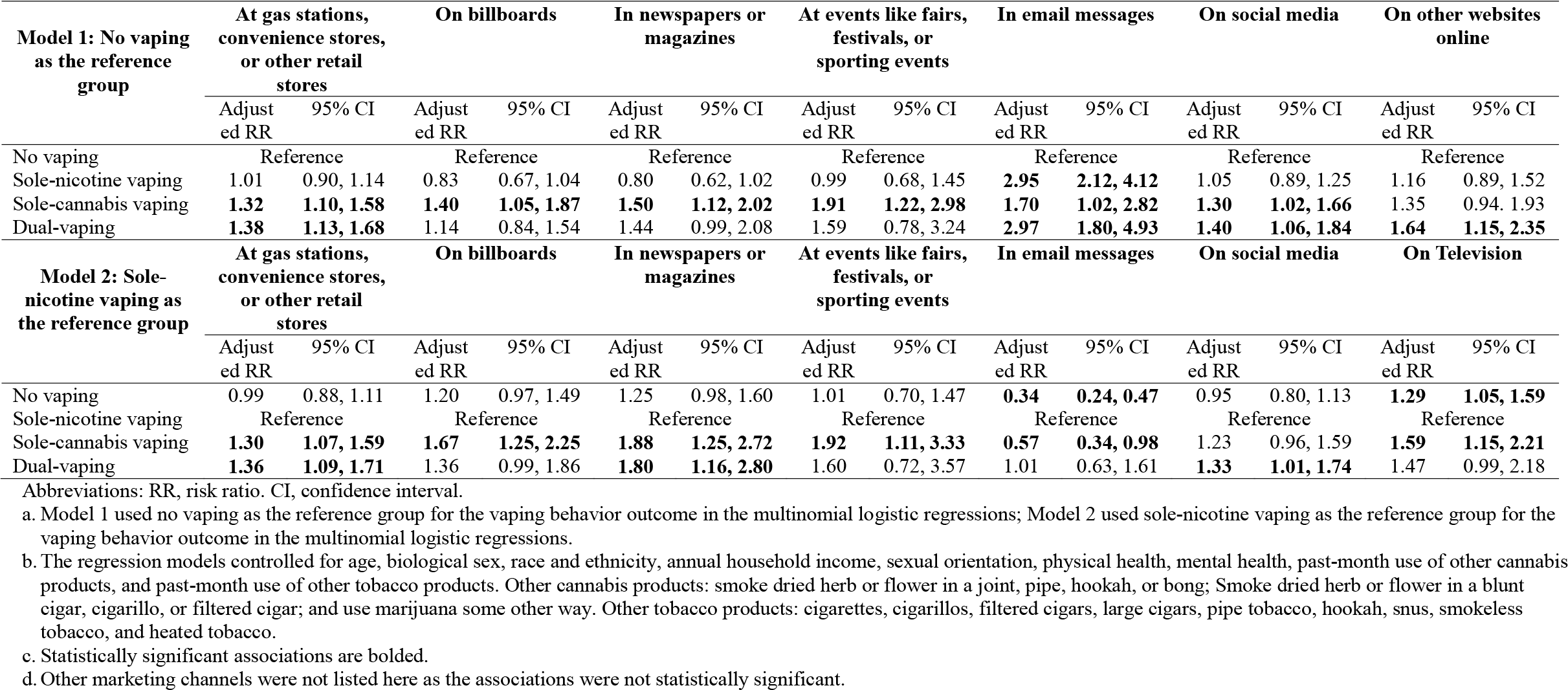
Associations between past 30-day e-cigarette marketing exposure (by marketing channels) and past 30-day vaping behavior^a-d^.

Using sole-nicotine vaping as the reference group (**Table 5**), adults who were exposed to e-cigarette marketing through five channels (retail stores; billboards; newspapers or magazines; events; television) had increased odds of sole-cannabis vaping (aRRs range between 1.30 and 1.92). Additionally, adults who were exposed to e-cigarette marketing through three channels (retail stores; newspapers or magazines; and social media) had increased odds of dual-vaping (aRRs range between 1.33 and 1.80).

**Supplemental Tables 2 and 3** represent additional results regarding associations between past 30-day e-cigarette marketing exposure (by marketing channels) and vaping behavior by age using both no vaping and sole-nicotine vaping as reference groups.

## DISCUSSION

Among a nationally representative sample of U.S. adults in 2021, those who were exposed (versus not exposed) to e-cigarette marketing were about 1.3-times more likely to report sole-cannabis vaping compared to those who reported no vaping or sole-nicotine vaping. Those exposed were also about 1.3-times more likely to report dual-vaping behaviors compared to those who reported no vaping. These were true even rigorously controlling for a number of socio-demographic, physical and mental health statuses, and substance use factors. Importantly, such associations appeared to be mainly driven by young adults aged 18-24 and 25-34 years, and no associations were found between e-cigarette marketing exposure and sole-nicotine vaping overall or among any age groups.

It is important to note that in this study, e-cigarette marketing could have been interpreted by respondents to include marketing for either or both nicotine and cannabis vape products. Due to the high resemblance of the product design and packages (including product shapes and sizes and product flavors) of nicotine and cannabis vape products,^25^ audiences may confuse ads for promoting these two vape products. This may be especially true as an increasing number of physical and online vape shops sell both nicotine and cannabis vape products,^26–29^ and in many cases, it is difficult to determine the ingredient in the vape products,^25^ and some vaping products can be used with nicotine and cannabis interchangeably.^26^ Therefore, e-cigarette marketing that promotes nicotine vaping products might contribute to vaping cannabis and vice versa. Additionally, more recently, multiple tobacco companies have started to invest in the cannabis industry, potentially using similar marketing tactics and materials to target the same communities and consumers to increase the sales of both nicotine and cannabis products.^30,31^

Our findings that e-cigarette marketing exposure was only associated with sole-cannabis vaping and dual-vaping behaviors, but not with sole-nicotine vaping, may be explained by dissimilar development of advertising restrictions for nicotine and cannabis products in recent years. First, various advertising restrictions and reduced advertising expenditures on nicotine vape products may have served to diminish the impact of e-cigarette marketing on nicotine vape product use. As the U.S. Food and Drug Administration (FDA) has tightened marketing regulations for nicotine vape products since 2019,^32^ the tobacco industry expenditure on e-cigarette marketing through various channels (e.g., print, television, websites) has declined significantly.^33^ Further, the FDA’s premarket tobacco product review process (which began in September 2020 for nicotine vape products) considers the influence of the new products’ marketing plans on increasing nicotine vaping among young people.^34^ Given these factors, the presence and impact of e-cigarette marketing on prompting nicotine vaping may have been measurably reduced prior to the administration of this Wave 6 survey in 2021. According to our study results, the only remaining e-cigarette marketing channel with a significant potential impact on nicotine vaping is email messaging, a growing direct-to-consumer marketing strategy to target future e-cigarette consumers and promote customer loyalty.^35,36^

By contrast, there are limited advertising restrictions for cannabis products, including for cannabis vape products. Given the current federal illegal status of cannabis, there are no federal-level cannabis marketing policies, and state and local-level policies related to cannabis advertising restrictions vary significantly.^37–39^ Further, compliance with such restrictions has been found to be low,^10,40–42^ and most restrictions do not apply to hemp-derived cannabis products which are rapidly gaining popularity.^43^ The lack of effective marketing restrictions on cannabis products may explain our finding that e-cigarette marketing exposure through many channels was positively associated with sole-cannabis vaping. In particular, marketing exposure through commercial events, including fairs or festivals, represented the strongest associations with sole-cannabis vaping. Unlike the nationwide restrictions on marketing tobacco products through commercial events,^44^ such restrictions do not exist for cannabis products, and many states allow cannabis industry-sponsored events as long as they are targeted to individuals “reasonably expected to be 21 years or older.”^45^ Therefore, continued monitoring and evaluation of cannabis marketing regulations at the local level is needed to understand their impact on cannabis consumption behaviors.

Our study results also showed that young adults aged 18-24 and 25-34 years have a higher prevalence of reporting sole-cannabis vaping and dual-vaping than older age groups, and significant associations between e-cigarette marketing exposure and sole-cannabis vaping were only found among those young adult age groups. These findings suggest e-cigarette marketing may be especially impactful for promoting cannabis vape product use among young adults. With the continued rise in cannabis vaping prevalence among young adults in the country,^2^ more research is needed to investigate the role of e-cigarette marketing, and more directly cannabis vape product marketing, on cannabis vaping behaviors among this population. For example, it may be important to examine the influence of specific marketing features (e.g., flavor descriptions, price promotions, human models, health claims, warnings)^46–49^ on cannabis vape product use perceptions and behaviors among young adults.

Among the limitations of this study are those related to the measures. The PATH Study assessment for e-cigarette marketing exposure asks about “e-cigarettes or other electronic nicotine products.” However, it is unclear how respondents interpreted and responded to the assessment (i.e., considering only nicotine vape products or also cannabis vape products). Therefore, research is needed to develop more specific measures that disentangle marketing exposure for nicotine versus cannabis vape products and to examine whether the impacts of e-cigarette marketing on vaping behavior show product use specificity or generalize across product categories. Further, measures to differentiate cannabis use motives (e.g., medical use or recreational use or both) or the various types of vaped cannabinoids (CBD vs. THC vs. others) were not included, which warrants consideration in future research regarding the impact of e-cigarette marketing exposure.^38,51^ Other limitations include the cross-sectional design, which limits causal inference, and small cell sizes limiting power (e.g., use among older age groups).

## CONCLUSIONS

This cross-sectional study highlights the associations between e-cigarette marketing exposure and sole-cannabis and dual-nicotine and cannabis vaping in U.S. adults, particularly among young adults aged 18-24 and 25-34 years. A striking finding is that sole-nicotine vaping was not associated with e-cigarette marketing exposure, suggesting that greater restrictions on tobacco marketing in recent years may have reduced the influence of e-cigarette marketing on nicotine vaping, while gaps in such marketing restrictions for cannabis may contribute to the influence of e-cigarette marketing on cannabis vaping. Continued monitoring and evaluation of the tobacco and cannabis industries’ marketing practices and their influence is critically needed to inform regulatory actions aimed at minimizing the public harm of nicotine and cannabis product use.

## Data Availability

All data produced are available online at https://www.icpsr.umich.edu/web/NAHDAP/studies/36498

