## Supplemental tables for "Associations between e-cigarette marketing exposure and vaping nicotine and cannabis among U.S. adults, 2021"

Supplemental Table 1. Weighted prevalence of past 30-day e-cigarette marketing exposure by marketing channel, age, and past 30-day vaping behavior^a^

| **Adults Aged 18-24 Years** | **All respondents** | | **No vaping** | | | **Sole-nicotine vaping** | | | **Sole-cannabis vaping** | | | **Dual-vaping** | |
| --- | --- | --- | --- | --- | --- | --- | --- | --- | --- | --- | --- | --- | --- |
|  | % | 95% CI | | % | 95% CI | | % | 95% CI | % | 95% CI | % | | 95% CI |
| Any marketing channel | 60.5 | 59.3, 61.7 | | 60.2 | 58.8, 61.6 | | 58.0 | 55.0, 61.0 | 71.0 | 66.4, 75.3 | 62.2 | | 57.0, 67.2 |
| At gas stations, convenience stores, or other retail stores | 49.2 | 47.9, 50.5 | | 48.4 | 46.9, 49.8 | | 47.3 | 44.1, 50.6 | 62.5 | 58.1, 66.7 | 53.4 | | 47.6, 59.0 |
| On billboards | 11.2 | 10.5, 11.8 | | 11.7 | 10.8, 12.5 | | 7.5 | 6.2, 9.1 | 13.8 | 10.6, 17.8 | 11.7 | | 8.7, 15.6 |
| In newspapers or magazines | 3.6 | 3.2, 4.1 | | 3.9 | 3.4, 4.4 | | 2.4 | 1.6, 3.5 | 4.6 | 3.0, 7.1 | 2.5 | | 1.4, 4.4 |
| On radio | 5.1 | 4.6, 5.6 | | 5.1 | 4.6, 5.7 | | 5.8 | 4.2, 7.9 | 3.9 | 2.5, 6.0 | 3.1 | | 1.7, 5.6 |
| On Television | 11.8 | 11.1, 12.5 | | 12.9 | 12.1, 13.7 | | 7.8 | 6.4, 9.5 | 8.5 | 6.2, 11.4 | 10.1 | | 7.1, 14.2 |
| At events like fairs, festivals, or sporting events | 1.8 | 1.6, 2.2 | | 1.8 | 1.5, 2.2 | | 1.5 | 0.9, 2.5 | 3.1 | 1.8, 5.4 | 1.7 | | 0.8, 3.2 |
| At nightclubs, bars, or music concerts | 3.7 | 3.3, 4.2 | | 3.5 | 3.1, 3.9 | | 3.0 | 2.0, 4.5 | 7.6 | 5.5, 10.4 | 5.2 | | 3.3, 8.3 |
| In email messages | 1.3 | 1.1, 1.6 | | 1.1 | 0.8, 1.3 | | 2.0 | 1.4, 3.0 | 2.3 | 1.4, 3.7 | 2.1 | | 1.2, 3.8 |
| On social media | 19.6 | 18.8, 20.5 | | 20.3 | 19.3, 21.3 | | 15.3 | 13.3, 17.6 | 22.4 | 18.1, 27.3 | 19.0 | | 15.7, 22.9 |
| On other websites online | 5.5 | 5.0, 6.1 | | 5.6 | 5.0, 6.3 | | 4.6 | 3.4, 6.1 | 6.5 | 4.3, 9.7 | 6.8 | | 4.8, 9.5 |
| Somewhere else | 0.5 | 0.4, 0.7 | | 0.6 | 0.4, 0.8 | | 0.3 | 0.1, 1.0 | 0.1 | 0.0, 0.6 | 0.8 | | 0.3, 2.4 |
| **Adults Aged 25-34 Years** | **All participants** | | **No vaping** | | | **Sole-nicotine vaping** | | | **Sole-cannabis vaping** | | | **Dual-vaping** | |
|  | % | 95% CI | | % | 95% CI | | % | 95% CI | % | 95% CI | % | | 95% CI |
| Any marketing channel | 58.2 | 56.6, 59.8 | | 57.1 | 55.2, 59.0 | | 59.5 | 55.8, 63.0 | 66.6 | 60.6, 72.1 | 66.8 | | 59.8, 73.2 |
| At gas stations, convenience stores, or other retail stores | 48.1 | 46.3, 49.8 | | 46.5 | 44.5, 48.6 | | 50.6 | 47.0, 54.5 | 58.4 | 52.3, 64.3 | 61.8 | | 54.7, 68.4 |
| On billboards | 9.8 | 8.8, 10.9 | | 9.4 | 8.3, 10.7 | | 10.2 | 7.4, 13.7 | 13.2 | 9.9, 17.2 | 13.8 | | 8.8, 21.0 |
| In newspapers or magazines | 5.5 | 4.8, 6.3 | | 5.0 | 4.2, 5.8 | | 6.6 | 4.7, 9.1 | 7.6 | 5.2, 11.2 | 11.1 | | 6.4, 18.7 |
| On radio | 5.5 | 4.9, 6.1 | | 5.4 | 4.8, 6.1 | | 4.8 | 3.3, 6.8 | 6.1 | 3.7, 10.0 | 9.1 | | 4.6, 17.3 |
| On Television | 10.1 | 9.1, 11.2 | | 9.9 | 8.8, 11.1 | | 10.2 | 7.8, 13.2 | 12.1 | 9.1, 15.9 | 13.1 | | 7.8, 21.3 |
| At events like fairs, festivals, or sporting events | 2.3 | 1.9, 2.8 | | 2.1 | 1.7, 2.7 | | 2.9 | 1.7, 5.0 | 3.0 | 1.7, 5.1 | 3.7 | | 1.0, 12.7 |
| At nightclubs, bars, or music concerts | 4.8 | 4.2, 5.4 | | 4.5 | 3.9, 5.2 | | 6.6 | 4.4, 9.9 | 5.0 | 3.1, 8.0 | 5.5 | | 3.0, 10.1 |
| In email messages | 2.1 | 1.7, 2.5 | | 1.5 | 1.1, 2.1 | | 5.2 | 3.7, 7.4 | 2.9 | 1.5, 5.5 | 5.0 | | 2.8, 8.9 |
| On social media | 15.7 | 14.4, 17.0 | | 15.4 | 13.9, 17.0 | | 16.2 | 13.2, 19.9 | 16.6 | 12.7, 21.4 | 20.1 | | 14.3, 27.5 |
| On other websites online | 5.2 | 4.4, 6.1 | | 5.0 | 4.1, 6.1 | | 5.6 | 3.6, 8.5 | 5.4 | 3.4, 8.4 | 8.0 | | 4.8, 13.0 |
| Somewhere else | 0.9 | 0.6, 1.3 | | 0.9 | 0.6, 1.5 | | 0.7 | 0.2, 2.5 | 0.3 | 0.1, 1.3 | 0.6 | | 0.1, 2.2 |
| **Adults Aged 35-54 Years** | **All respondents** | | **No vaping** | | | **Sole-nicotine vaping** | | | **Sole-cannabis vaping** | | | **Dual-vaping** | |
|  | % | 95% CI | | % | 95% CI | | % | 95% CI | % | 95% CI | % | | 95% CI |
| Any marketing channel | 54.0 | 52.2, 55.8 | | 53.5 | 51.5, 55.4 | | 57.0 | 52.0, 61.9 | 62.0 | 53.8, 69.5 | 67.5 | | 51.6, 80.2 |
| At gas stations, convenience stores, or other retail stores | 45.1 | 43.2, 47.0 | | 44.5 | 42.5, 46.6 | | 48.4 | 43.1, 53.8 | 53.2 | 45.1, 61.2 | 59.6 | | 46.4, 71.5 |
| On billboards | 8.3 | 7.4, 9.4 | | 8.2 | 7.2, 9.4 | | 8.7 | 5.8, 12.8 | 11.3 | 7.0, 17.7 | 4.7 | | 1.9, 11.1 |
| In newspapers or magazines | 5.9 | 5.0, 6.8 | | 5.9 | 5.0, 6.9 | | 4.6 | 2.8, 7.5 | 8.1 | 5.2, 12.5 | 4.9 | | 2.1, 11.4 |
| On radio | 6.2 | 5.3, 7.3 | | 6.1 | 5.2, 7.2 | | 5.2 | 3.0, 8.7 | 10.7 | 6.7, 16.7 | 4.6 | | 1.8, 11.0 |
| On Television | 11.7 | 10.6, 12.9 | | 11.8 | 10.6, 13.1 | | 9.1 | 6.3, 13.0 | 13.1 | 8.6, 19.4 | 11.1 | | 5.8, 20.4 |
| At events like fairs, festivals, or sporting events | 1.7 | 1.2, 2.3 | | 1.6 | 1.1, 2.3 | | 1.3 | 0.5, 3.3 | 4.0 | 1.8, 8.6 | 3.6 | | 1.1, 11.6 |
| At nightclubs, bars, or music concerts | 2.6 | 2.1, 3.2 | | 2.5 | 2.0, 3.1 | | 3.6 | 1.8, 6.8 | 3.5 | 1.5, 7.7 | 5.7 | | 2.5, 12.2 |
| In email messages | 1.1 | 0.9, 1.4 | | 1.0 | 0.7, 1.3 | | 3.7 | 2.3, 6.1 | 1.1 | 0.4, 3.5 | 4.3 | | 1.8, 9.7 |
| On social media | 8.9 | 8.0, 9.8 | | 8.5 | 7.6, 9.5 | | 11.3 | 8.1, 15.6 | 13.4 | 9.1, 19.1 | 14.8 | | 7.7, 26.6 |
| On other websites online | 3.6 | 3.0, 4.4 | | 3.5 | 2.8, 4.3 | | 4.8 | 3.0, 7.7 | 5.8 | 3.1, 10.5 | 6.1 | | 2.6, 13.8 |
| Somewhere else | 0.6 | 0.4, 0.9 | | 0.6 | 0.4, 0.9 | | 0.8 | 0.4, 1.5 | 0.5 | 0.1, 3.4 | NA | | NA |
| **Adults Aged ≥55 Years** | **All respondents** | | **No vaping** | | | **Sole-nicotine vaping** | | | **Sole-cannabis vaping** | | | **Dual-vaping** | |
|  | % | 95% CI | | % | 95% CI | | % | 95% CI | % | 95% CI | % | | 95% CI |
| Any marketing channel | 45.3 | 43.5, 47.1 | | 45.1 | 43.3, 47.0 | | 49.6 | 40.6, 58.6 | 53.4 | 40.5, 65.8 | 62.0 | | 36.5, 82.2 |
| At gas stations, convenience stores, or other retail stores | 35.9 | 34.0, 37.8 | | 35.7 | 33.8, 37.7 | | 37.5 | 29.2, 46.6 | 44.7 | 33.6, 56.3 | 62.0 | | 36.5, 82.2 |
| On billboards | 8.0 | 7.0, 9.1 | | 8.0 | 7.0, 9.1 | | 4.3 | 2.1, 8.8 | 10.6 | 5.2, 20.5 | 12.7 | | 3.9, 34.2 |
| In newspapers or magazines | 7.2 | 6.3, 8.3 | | 7.2 | 6.3, 8.3 | | 2.5 | 0.9, 6.7 | 11.4 | 5.2, 22.9 | 13.4 | | 4.4, 34.3 |
| On radio | 3.2 | 2.7, 3.7 | | 3.0 | 2.6, 3.6 | | 9.2 | 4.0, 19.9 | 2.9 | 1.4, 5.8 | 9.3 | | 2.4, 29.8 |
| On Television | 10.4 | 9.3, 11.7 | | 10.5 | 9.3, 11.8 | | 7.0 | 4.2, 11.4 | 9.8 | 4.1, 21.5 | 9.6 | | 2.6, 29.7 |
| At events like fairs, festivals, or sporting events | 1.5 | 1.2, 1.9 | | 1.5 | 1.1, 1.9 | | 1.0 | 0.2, 4.2 | 2.8 | 1.3, 5.9 | NA | | NA |
| At nightclubs, bars, or music concerts | 1.6 | 1.2, 2.1 | | 1.6 | 1.2, 2.2 | | 2.6 | 0.9, 7.0 | 1.0 | 0.2, 4.0 | NA | | NA |
| In email messages | 0.8 | 0.6, 1.0 | | 0.7 | 0.5, 1.0 | | 4.0 | 1.5, 10.4 | 1.3 | 0.3, 5.1 | 9.3 | | 2.4, 29.8 |
| On social media | 4.1 | 3.5, 4.8 | | 4.1 | 3.5, 4.8 | | 2.1 | 0.9, 5.0 | 4.2 | 1.1, 14.7 | 11.7 | | 2.3, 42.5 |
| On other websites online | 1.5 | 1.1, 1.9 | | 1.5 | 1.1, 1.9 | | 1.8 | 0.7, 4.6 | 2.4 | 1.0, 5.5 | NA | | NA |

Abbreviations: NA, not available due to small sample estimates; CI, confidence interval.

a. All percentages are weighted.

Supplemental Table 2. Associations between past 30-day e-cigarette marketing exposure (by marketing channel) and vaping behavior by age using no vaping as the reference group^a-d^

| **Adults Aged 18-24 Years** | **At gas stations, convenience stores, or other retail stores** | | **On billboards** | | | **In newspapers or magazines** | | **On Television** | | | **At nightclubs, bars, or music concerts** | | **In email messages** | | **On social media** | |
| --- | --- | --- | --- | --- | --- | --- | --- | --- | --- | --- | --- | --- | --- | --- | --- | --- |
|  | Adjusted RR | 95% CI | Adjusted RR | | 95% CI | Adjusted RR | 95% CI | Adjusted RR | | 95% CI | Adjusted RR | 95% CI | Adjusted RR | 95% CI | Adjusted RR | 95% CI |
| No vaping | Reference | | Reference | | | Reference | | Reference | | | Reference | | Reference | | Reference | |
| Sole-nicotine vaping | 0.89 | 0.77, 1.04 | **0.62** | **0.48, 0.79** | | **0.62** | **0.39, 0.99** | **0.62** | **0.47, 0.81** | | 0.78 | 0.49, 1.25 | **1.97** | **1.18, 3.30** | **0.77** | **0.63, 0.93** |
| Sole-cannabis vaping | **1.58** | **1.29, 1.93** | 1.11 | 0.78, 1.58 | | 1.17 | 0.74, 1.86 | 0.77 | 0.54, 1.10 | | **1.79** | **1.17, 2.74** | **2.29** | **1.26, 4.17** | 1.19 | 0.89, 1.59 |
| Dual-vaping | 1.03 | 0.80, 1.33 | 0.97 | 0.65, 1.45 | | 0.66 | 0.32, 1.35 | 0.93 | 0.60, 1.44 | | 1.22 | 0.73, 2.02 | **2.30** | **1.15, 4.59** | 1.04 | 0.79, 1.37 |
| **Adults Aged 25-34 Years** | **At gas stations, convenience stores, or other retail stores** | | **In newspapers or magazines** | | | **On Television** | | **In email messages** | | | **On other websites online** | |  | |  | |
|  | Adjusted RR | 95% CI | Adjusted RR | 95% CI | | Adjusted RR | 95% CI | Adjusted RR | 95% CI | | Adjusted RR | 95% CI |  |  |  |  |
| No vaping | Reference | | Reference | | | Reference | | Reference | | | Reference | |  | |  | |
| Sole-nicotine vaping | 1.16 | 0.97, 1.39 | 1.43 | 0.91, 2.23 | | 1.09 | 0.75, 1.57 | **2.88** | **1.70, 4.88** | | 1.27 | 0.77, 2.10 |  |  |  |  |
| Sole-cannabis vaping | **1.36** | **1.03, 1.80** | 1.64 | 0.99, 2.68 | | **1.61** | **1.05, 2.46** | 1.77 | 0.71, 4.42 | | 1.22 | 0.68, 2.18 |  |  |  |  |
| Dual-vaping | **1.61** | **1.15, 2.26** | **2.45** | **1.33, 4.51** | | 1.73 | 0.89, 3.35 | 2.49 | 0.98, 6.35 | | **1.94** | **1.03, 3.66** |  |  |  |  |
| **Adults Aged 35-54 Years** | **On radio** | | **At events like fairs, festivals, or sporting events** | | | **In email messages** | | **On social media** | | |  | |  | |  |  |
|  | Adjusted RR | 95% CI | Adjusted RR | 95% CI | | Adjusted RR | 95% CI | Adjusted RR | 95% CI | |  |  |  |  |  |  |
| No vaping | Reference | | Reference | | | Reference | | Reference | | |  | |  | |  |  |
| Sole-nicotine vaping | 0.86 | 0.45, 1.64 | 0.78 | 0.22, 2.76 | | **3.32** | **1.64, 6.71** | **1.53** | **1.01, 2.29** | |  |  |  |  |  |  |
| Sole-cannabis vaping | **1.93** | **1.03, 3.61** | **2.76** | **1.12, 6.83** | | 1.03 | 0.26, 4.11 | **1.90** | **1.16, 3.11** | |  |  |  |  |  |  |
| Dual-vaping | 0.70 | 0.24, 2.01 | 2.45 | 0.49, 9.30 | | 3.20 | 0.92, 9.18 | 1.79 | 0.74, 4.33 | |  |  |  |  |  |  |

Abbreviations: RR, risk ratio. CI, confidence interval.

a. The regression models controlled for age, biological sex, race and ethnicity, annual household income, sexual orientation, physical health, mental health, past-month use of other cannabis products, and past-month use of other tobacco products. Other cannabis products: smoke dried herb or flower in a joint, pipe, hookah, or bong; Smoke dried herb or flower in a blunt cigar, cigarillo, or filtered cigar; and use marijuana some other way. Other tobacco products: cigarettes, cigarillos, filtered cigars, large cigars, pipe tobacco, hookah, snus, smokeless tobacco, and heated tobacco.
b. Statistically significant associations are bolded.
c. Results from the regression models for those aged ≥55 years were omitted due to model convergence issues from small cell size.
d. Other channels were not listed here as the associations were not statistically significant.

Supplemental Table 3. Associations between past 30-day e-cigarette marketing exposure (by marketing channel) and past 30-day vaping behavior by age using sole-nicotine vaping as the reference group^a-d^

| **Adults Aged 18-24 Years** | **At gas stations, convenience stores, or other retail stores** | | | **On billboards** | | | **In newspapers or magazines** | | **On television** | | **At nightclubs, bars, or music concerts** | | **In email messages** | | **On social media** | |
| --- | --- | --- | --- | --- | --- | --- | --- | --- | --- | --- | --- | --- | --- | --- | --- | --- |
|  | Adjusted RR | 95% CI | Adjusted RR | | 95% CI | Adjusted RR | | 95% CI | Adjusted RR | 95% CI | Adjusted RR | 95% CI | Adjusted RR | 95% CI | Adjusted RR | 95% CI |
| No vaping | 1.12 | 0.96, 1.30 | **1.62** | | **1.27, 2.07** | **1.61** | | **1.01, 2.57** | **1.61** | **1.23, 2.11** | 1.28 | 0.80, 2.04 | **0.51** | **0.30, 0.85** | **1.30** | **1.07, 1.59** |
| Sole-nicotine vaping | Reference | | | Reference | | | Reference | | Reference | | Reference | | Reference | | Reference | |
| Sole-cannabis vaping | **1.76** | **1.39, 2.24** | 1.80 | | 1.25, 2.58 | 1.89 | | 0.99, 3.55 | 1.24 | 0.83, 1.85 | **2.29** | **1.31, 4.01** | 1.16 | 0.56, 2.41 | **1.56** | **1.11, 2.17** |
| Dual-vaping | 1.15 | 0.87, 1.52 | 1.58 | | 0.99, 2.52 | 1.06 | | 0.54, 2.08 | 1.50 | 0.94, 2.40 | 1.55 | 0.82, 2.93 | 1.17 | 0.55, 2.48 | **1.35** | **1.01, 1.81** |
| **Adults Aged 25-34 Years** | **In email messages** | | |  | | |  | |  | |  | |  | |  | |
|  | Adjusted RR | 95% CI |  | |  |  | |  |  |  |  |  |  |  |  |  |
| No vaping | **0.35** | **0.20, 0.59** |  | |  |  | |  |  |  |  |  |  |  |  |  |
| Sole-nicotine vaping | Reference | | |  | | |  | |  | |  | |  | |  | |
| Sole-cannabis vaping | 0.61 | 0.26, 1.43 |  | |  |  | |  |  |  |  |  |  |  |  |  |
| Dual-vaping | 0.86 | 0.37, 2.00 |  | |  |  | |  |  |  |  |  |  |  |  |  |
| **Adults Aged 35-54 Years** | **In newspapers or magazines** | | | **On radio** | | | **In email messages** | | **On social media** | |  | |  | |  | |
|  | Adjusted RR | 95% CI | Adjusted RR | | 95% CI | Adjusted RR | | 95% CI | Adjusted RR | 95% CI |  |  |  |  |  |  |
| No vaping | 1.50 | 0.85, 2.63 | 1.16 | | 0.61, 2.21 | **0.30** | | **0.15, 0.61** | **0.66** | **0.44, 0.99** |  |  |  |  |  |  |
| Sole-nicotine vaping | Reference | | | Reference | | | Reference | | Reference | |  | |  | |  | |
| Sole-cannabis vaping | **2.49** | **1.16, 5.33** | **2.24** | | **1.01, 4.96** | 0.31 | | 0.07, 1.45 | 1.24 | 0.68, 2.28 |  |  |  |  |  |  |
| Dual-vaping | 1.34 | 0.44, 4.08 | 0.81 | | 0.24, 2.73 | 0.97 | | 0.27, 3.50 | 1.18 | 0.48, 2.88 |  |  |  |  |  |  |

Abbreviations: RR, risk ratio. CI, confidence interval.

e. The regression models controlled for age, biological sex, race and ethnicity, annual household income, sexual orientation, physical health, mental health, past 30-day use of other cannabis products, and past 30-day use of other tobacco products. Other cannabis products: smoke dried herb or flower in a joint, pipe, hookah, or bong; Smoke dried herb or flower in a blunt cigar, cigarillo, or filtered cigar; and use marijuana some other way. Other tobacco products: cigarettes, cigarillos, filtered cigars, large cigars, pipe tobacco, hookah, snus, smokeless tobacco, and heated tobacco.
f. Statistically significant associations are bolded.
g. Results from the regression models for those aged ≥55 years were omitted due to model convergence issues from small cell size.
h. Other channels were not listed here as the associations were not statistically significant.
